# Increased serum levels of sCD14 and sCD163 indicate a preponderant role for monocytes in COVID-19 immunopathology

**DOI:** 10.1101/2020.06.02.20120295

**Authors:** J Gómez-Rial, MJ Currás-Tuala, I Rivero-Calle, A Gómez-Carballa, M Cebey-López, C Rodríguez-Tenreiro, A Dacosta-Urbieta, C Rivero-Velasco, N Rodríguez-Núñez, R Trastoy-Pena, J Rodríguez-García, A Salas, F Martinón-Torres, on behalf of the GEN-COVID Study Group

**Author notes:** **Correspondence:** Dr Federico Martinón-Torres. Equally contributed to this manuscript (J Gómez-Rial, MJ Currás-Tuala). GEN-COVID Study Group (www.gencovid.es): Aguilera Guirao, Antonio^3^; Álvarez Escudero, Julián^7^; Antela López, Antonio^5^; Barbeito Castiñeiras, Gema^3^; Bello Paderne, Xabier^1^; Ben García, Miriam^1^; Carral García, María Victoria^12^; Cebey López, Miriam^1^; Coira Nieto, Amparo^3^; Conde Pájaro, Mónica^9^; Costa Alcalde, José Javier^3^; Currás Tuala, María José^1^; Dacosta Urbieta, Ana Isabel^1^; Díaz Esteban, Blanca^1^; Domínguez Santalla, María Jesús^5^; Fernández Villaverde, Juan^6^; Galbán Rodríguez, Cristóbal^6^; García Allut, José Luis^6^; García Vicente, Luisa^1^; Giráldez Vázquez, Elena^6^; Gómez Carballa, Alberto^1^; Gómez Rial, José^1^; González Barcala, Francisco Javier^4^; Guerra Liñares, Beatriz^9^; Leboráns Iglesias, Pilar^1^; Lence Massa, Beatriz^6^; Lendoiro Fuentes, Marta^1^; López Franco, Montserrat^1^; López Lago, Ana^6^; Navarro De la Cruz, Daniel^3^; Núñez Masid, Eloína^10^; Ortolá Devesa, Juan Bautista^8^; Pardo Seco, Jacobo^1^; Pazo Núñez, María^5^; Pérez del Molino Bernal, Marisa^3^; Pérez Freixo, Hugo^9^; Piñeiro Rodríguez, Lidia^1^; Pischedda, Sara^1^; Portela Romero, Manuel^11^; Pose Reino, Antonio^5^; Prada Hervella, Gloria María^7^; Queiro Verdes, Teresa^9^; Redondo Collazo, Lorenzo^1^; Regueiro Casuso, Patricia^1^; Rey García, Susana^1^; Rey Vázquez, Sara^1^; Riveiro Blanco, Vanessa^4^; Rivero Calle, Irene^1^; Rivero Velasco, Carmen^6^; Rodríguez Núñez, Nuria^4^; Rodríguez-Tenreiro Sánchez, Carmen^1^; Saborido Paz, Eva^6^; Sadiki Orayyou, José Miguel^1^; Saito Villanueva, Carla^6^; Serén Fernández, Sonia^1^; Souto Sanmartín, Pablo^9^; Taboada Muñiz, Manuel^7^; Trastoy Pena, Rocío^3^; Treviño Castellano, Mercedes^3^; Valdés Cuadrado, Luis^4^; Varela García, Pablo^5^; Vilas Iglesias, María Soledad^1^; Viz Lasheras, Sandra^1^. Translational Pediatrics and Infectious Diseases, Pediatrics Department, Hospital Clínico Universitario de Santiago, Santiago de Compostela, Spain, and GENVIP Research Group (www.genvip.org), Instituto de Investigación Sanitaria de Santiago, Universidad de Santiago de Compostela, Galicia, Spain. Unidade de Xenética, Departamento de Anatomía Patolóxica e Ciencias Forenses, Instituto de Ciencias Forenses, Facultade de Medicina, Universidade de Santiago de Compostela, and GenPop Research Group, Instituto de Investigaciones Sanitarias (IDIS), Hospital Clínico Universitario de Santiago, Galicia, Spain. Microbiology Service, Hospital Clínico Universitario de Santiago, Santiago de Compostela, Galicia, Spain. Pneumology Service, Hospital Clínico Universitario de Santiago, Santiago de Compostela, Galicia, Spain. Internal Medicine Service, Hospital Clínico Universitario de Santiago, Santiago de Compostela, Galicia, Spain. Intensive Care Service, Hospital Clínico Universitario de Santiago, Santiago de Compostela, Galicia, Spain. Anesthesiology and Resuscitation Service, Hospital Clínico Universitario de Santiago, Santiago de Compostela, Galicia, Spain. Clinical Chemistry Laboratory, Hospital Clínico Universitario de Santiago, Santiago de Compostela, Galicia, Spain. Preventive Medicine Unit, Hospital Clínico Universitario de Santiago, Santiago de Compostela, Galicia, Spain. Manager of the Health Care Area of Santiago de Compostela and Barbanza, Hospital Clínico Universitario de Santiago, Santiago de Compostela, Galicia, Spain. Deputy Director of Pharmaceutical Delivery, Training, Teaching, Research and Innovation, Hospital Clínico Universitario de Santiago, Santiago de Compostela, Galicia, Spain. Director of Nursing Processes, Hospital Clínico Universitario de Santiago, Santiago de Compostela, Galicia, Spain.

## Abstract

**Background:** Emerging evidence indicates a potential role for monocyte in COVID-19 immunopathology. We investigated two soluble markers of monocyte activation, sCD14 and sCD163, in covid19 patients with the aim of characterizing their potential role in monocyte-macrophage disease immunopathology. To the best of our knowledge, this is the first study of its kind.

**Methods:** Fifty-nine SARS-Cov-2 positive hospitalized patients, classified according to ICU or non-ICU admission requirement, were prospectively recruited and analyzed by ELISA for levels of sCD14 and sCD163, along with other laboratory parameters, and compared to a healthy control group.

**Results:** sCD14 and sCD163 levels were significantly higher among COVID-19 patients, independently of ICU admission requirement, compared to the control group. We found a significant correlation between sCD14 levels and other inflammatory markers, particularly Interleukin-6, in the non-ICU patients’ group. sCD163 showed a moderate positive correlation with the time at sampling from admission, increasing its value over time, independently of severity group.

**Conclusions:** Monocyte-macrophage activation markers are increased and correlate with other inflammatory markers in SARS-Cov-2 infection, in association to hospital admission. These data suggest a potentially preponderant role for monocyte-macrophage activation in the development of immunopathology of covid19 patients.

## Introduction

Emerging evidence from SARS-Cov-2 infected patients suggests a key role for monocyte-macrophage in the immunopathology of COVID-19 infection, with a predominant monocyte-derived macrophage infiltration observed in severely damaged lungs [1], and morphological and inflammation-related changes in peripheral blood monocytes that correlate with the patients’ outcome [2] An overexuberant inflammatory immune response with production of a cytokine storm and T-cell immunosuppression are the main hallmarks of severity in these patients [3]. This clinical course resembles viral-associated hemophagocytic syndrome (VASH), a rare severe complication of various viral infections mediated by proinflammatory cytokines, resulting in multiorgan failure and death [4]. A chronic expansion of inflammatory monocytes and over-activation of macrophages have been extensively described in this syndrome [5; 6; 7]. VAHS has been identifies as a major contributor to death of patients in past pandemics outbreaks [8] including previous SARS and MERS outbreaks [9] and is currently suggested for SARS-Cov-2 outbreak. [10]

CD14 and CD163 are both myeloid differentiation markers found primarily on monocytes and macrophages, and detection of soluble release of both in plasma is considered a good biomarker of monocyte-macrophage activation [11; 12]. Elevated plasma levels of soluble CD14 (sCD14) are associated to poor prognosis in VIH-infected patients, are a strong predictor of morbidity and mortality [13; 14], and associated with diminished CD4+-T cell restoration [15]. In addition, soluble CD163 (sCD163) plasma levels are a good proxy for monocyte expansion and disease progression during HIV infection [16]. In measles infection, a leading cause of death associated with increased susceptibility to secondary infections and immunosuppression, sCD14 and sCD163 levels were found to be significantly higher, indicating an important and persistent monocyte-macrophage activation [17].

We hypothesized that monocytes/macrophages may be an important component of immunopathology associated to SARS-Cov-2 infection. In this paper, we analyze plasma levels of soluble monocyte activation markers in COVID-19 patients and their correlation with severity and other inflammatory markers.

## Methods

### Subjects

We recruited 59 patients with confirmed PCR-positive diagnostic for SARS-Cov-2 infection, classified according ICU admission requirement (n=22 patients), or non-ICU requirement (n=37), and age-matched healthy individuals (n=20) as control group. Demographic data, main medication treatment and routine lab clinical parameters including inflammatory biomarkers were collected for all infected patients. Leftover sera samples from routine analytical control were employed for the research analysis, after obtaining corresponding informed consent. Time elapsed from hospital admission to sample extraction was also recorded.

### Measurement of sCD14 and sCD163 serum levels

To determine levels of soluble monocyte activation markers in serum specimens, appropriate sandwich ELISA (Quantikine, R&D systems, United Kingdom) were used following manufacturer indications. Briefly, diluted sera samples were incubated for 3 hours at room temperature in the corresponding microplate strips coated with capture antibody. After incubation, strips were properly washed and incubated with the corresponding Human Antibody conjugate for 1 hour. After washing, reactions were revealed and optical density at 450 nm was determined in a microplate reader. Concentration levels were interpolated from the standard curve using a four-parameter logistic (4-PL) curve-fit in Prism8 GraphPad software. Final values were corrected applying the corresponding dilution factor employed.

### Statistical analysis

Data are expressed as median and interquartile range. All statistical analyses were performed using the statistical package R. Mann-Whitney tests were used for comparison between ICU and non-ICU groups *versus* healthy controls. Pearson’s correlation coefficients were used to quantify the association between sCD14 and sCD163 concentration and other lab parameters in non-ICU patients. Data outliers, falling outside the 1.5 interquartile range, were excluded from the statistical analysis. The nominal significance level considered was 0.05. Bonferroni adjustment was used to account for multiple testing.

## Results

### Demographic and clinical laboratory parameters

Patients in the ICU group showed significant differences in several clinical laboratory parameters when compared to non-ICU group: lymphocytes, ferritin, D-dimer, Lactate Dehydrogenase (LDH), Procalcitonin (PCT), Interleukin-6 (IL-6). The absolute value for circulating monocytes did not show significant differences between groups. However, these values may have been distorted by the use of tocilizumab, an IL-6 blocking drug extensively employed in the ICU group which interferes with monocyte function. Age and time elapsed from admission to sample extraction did not show differences between groups. Values are summarized in **Table 1**.

**Table 1.** Demographic and clinical laboratory parameters of patients recruited.

| Parameter | ICU | non-ICU | P-value |
| --- | --- | --- | --- |
| *Clinical laboratory parameters* |  |  |  |
| Lymphocytes | 0.54 (0.47–1.058) | 1.16 (0.79–1.62) | **0.0004** |
| Monocytes | 0.35 (0.16–0.65) | 0,42 (0.35–0.58) | ns |
| Platelets | 264 (204.3–354.5) | 272 (213–413) | ns |
| D-Dimer | 3676 (1198–8121) | 755 (413–1033) | **0.0002** |
| Lactate Dehydrogenase (LDH) | 677 (429–818.5) | 469 (391–595) | **0.0188** |
| C-reactive protein (CRP) | 7.37 (2.56–20.51) | 4,65 (2.16–11.41) | ns |
| Procalcitonin (PCT) | 0.22 (0.09–0.4) | 0.09 (0.05–0.21) | **0.0305** |
| Ferritin | 1257 (837.3–3020) | 467 (254.5–785) | **<0.0001** |
| Interleukin-6 (IL-6) | 83.10 (14.45–381.8) | 12.70 (6.95–46) | **0.0014** |
| Glycosylated Hemoglobin (Hb1Ac) | 5.95 (5.65–6.47) | 6.1 (5.7–6.9) | ns |
| Troponin-I | 0.021 (0.017–0.246) | 0.017 (0.017–0.019) | ns |
| *Time elapsed from admission to sample (days)* |  |  |  |
|  | 5 (3.75–10) | 4 (2–6) | ns |
| *Age (years)* |  |  |  |
|  | 52 (48.75–61.25) | 52 (44–65) | ns |
| *Tocilizumab/Corticoids* |  |  |  |
|  | 19/22 (87%) | 2/37 (5.4%) | **<0.0001** |

### Serum levels for sCD14 and sCD163

Median levels for sCD14 in sera from ICU patients were 2444.0 (95%CI: 1914.0-3251.0) ng/ml, compared to 2613.0 (95%CI: 2266.0-2991.0) ng/ml in non-ICU patients. The healthy control group median value was 1788.0 (95%CI: 1615.0-1917.0) ng/ml. We observed significant differences for values from infected patients relative to control group (*P*-value<0.0001), however no significant differences were observed between ICU and non-ICU groups. Median levels for sCD163 in sera from ICU patients were 911.5 (95%CI: 624.7-1167.0) ng/ml, and 910.4 (95%CI: 733.1-1088.0) ng/ml in non-ICU patients. The healthy control group value was 495.6 (95%CI: 332.5-600.7) ng/ml. In the same way as with sCD14, we observed significant differences for values from infected patients compared to control group (P-value<00001), but no differences between ICU and non-ICU infected patients. Values are summarized in **Table 2** and **Figure 1**.

**Table 2.** Concentration (ng/ml) of serum levels of sCD14 and sCD163 in patients from ICU and non-ICU groups, and healthy controls. Data are represented as median and interquartile range.

| **Concentration** | **ICU** | **non-ICU** | **Healthy controls** |
| --- | --- | --- | --- |
| sCD14 | 2444.0 (1914.0–3251.0) | 2613.0 (2266.0–2991.0) | 1788.0 (1615.0–1917.0) |
| sCD163 | 911.5 (624.7–1167) | 910.4 (733.1–1088) | 495.6 (332.5–600.7) |

**Figure 1.**
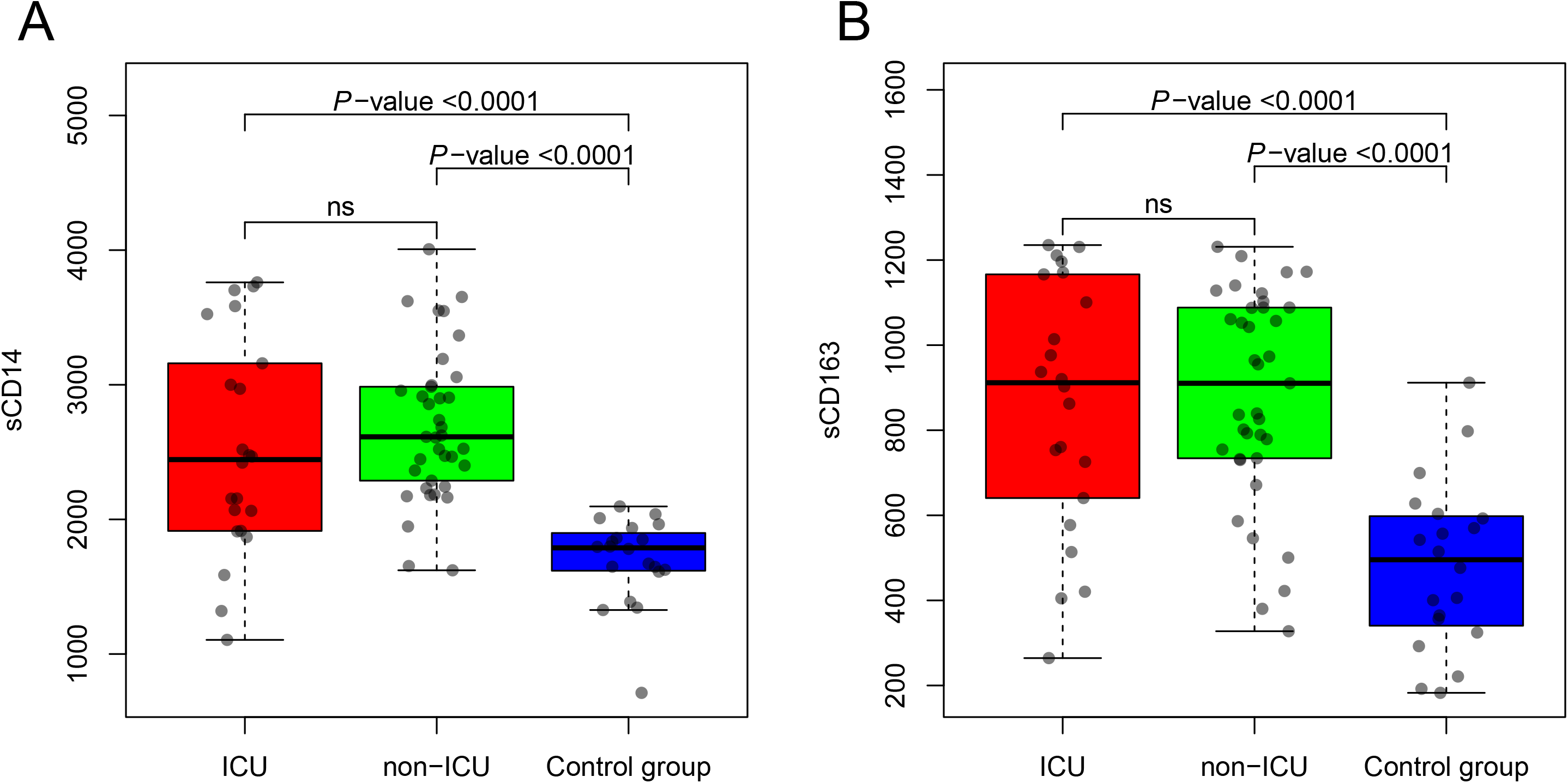
Values of sCD14 and sCD163 in sera samples from patients in ICU, non-ICU groups, and healthy controls. Results are presented as median and interquartile range levels in ng/ml. Non-parametric Mann-Whitney tests were used for comparison between groups, and P-values for the different comparisons are displayed.

### Correlation between sCD14 and sCD163 levels and time elapsed from hospital admission

We assessed correlation between sCD14 and sCD163 levels and time elapsed from hospital admission to sample extraction (**Figure 2**). We found a significant positive correlation between sCD163 levels and time elapsed (r^2^=0.3246, *P*-value=0.0156), increasing its value over time of hospital admission. We did not observe a significant correlation between sCD14 levels and time elapsed from hospital admission to sample extraction.

**Figure 2.**
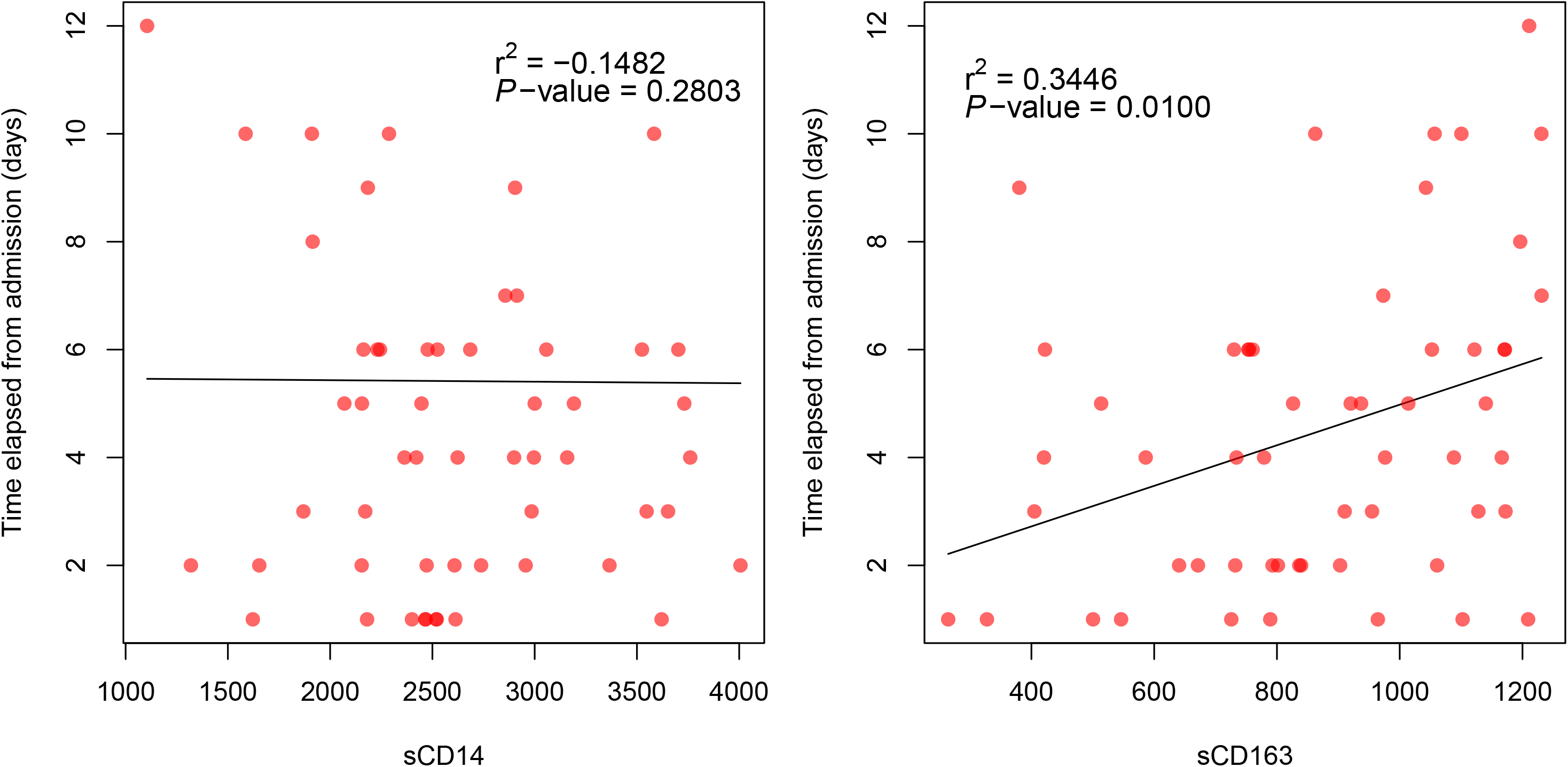
Correlation between serum levels of sCD14 and sCD163 and time elapsed from admission to sample extraction in days for all infected patients. Pearson’s correlation coefficient (r^2^) and *P*-value are shown.

### Correlation between sCD14 and sCD163 levels and clinical laboratory parameters

We found significant correlation between sCD14 and sCD163 and several clinical laboratory parameters in infected patients (in these analysis, adjusted significance under Bonferrori correction is 0.01), but only in the non-ICU group, possibly reflecting an interference of the use of tocilizumab or corticoids in the ICU group. Levels of sCD14 showed a negative correlation with the absolute value of lymphocytes (r^2^=-0.5501, P-value=0.0005) and a positive correlation with levels of LDH (r^2^=0.5906, P-value=0.0001), CRP (r^2^=0.6275, P-value<0.0001); PCT (r^2^=0.4608, P-value=0.0091), and Ferritin (r^2^=0.4414, P-value=0.0090) (**Figure 3**). No other significative associations were found with other lab parameters. Levels of sCD163 did not show any significant correlation with clinical laboratory parameters (**Figure 3**). Particularly, IL-6 also shows a significant positive correlation with sCD14 (r^2^=0.6034, P-value=0.0003) (**Figure 4**).

**Figure 3.**
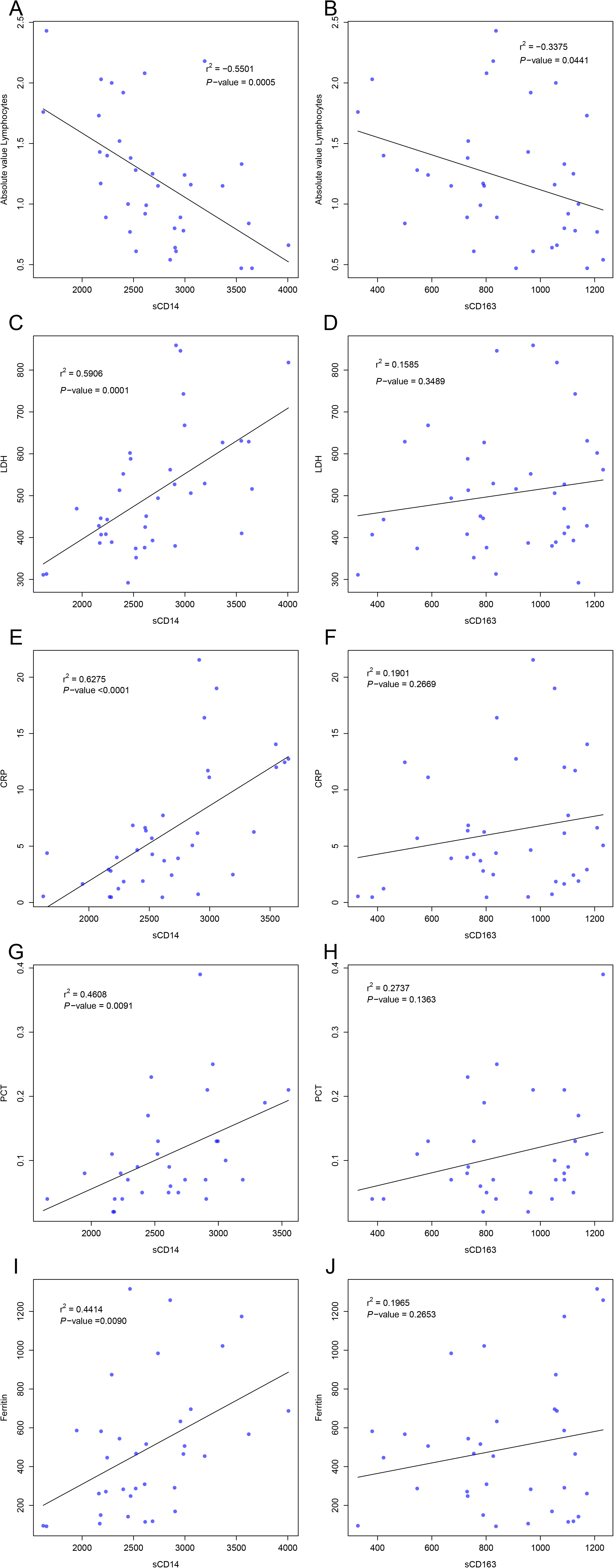
Association between serum levels of sCD14 and sCD163, and several laboratory parameters including Absolute Valor Lymphocytes, LDH, CRP, PCT and Ferritin in the non-ICU patient group. Pearson’s correlation coefficient (r^2^) and P-value are shown. LDH: Lactate Dehydrogenase; CRP: C-reactive Protein; PCT: Procalcitonin

**Figure 4.**
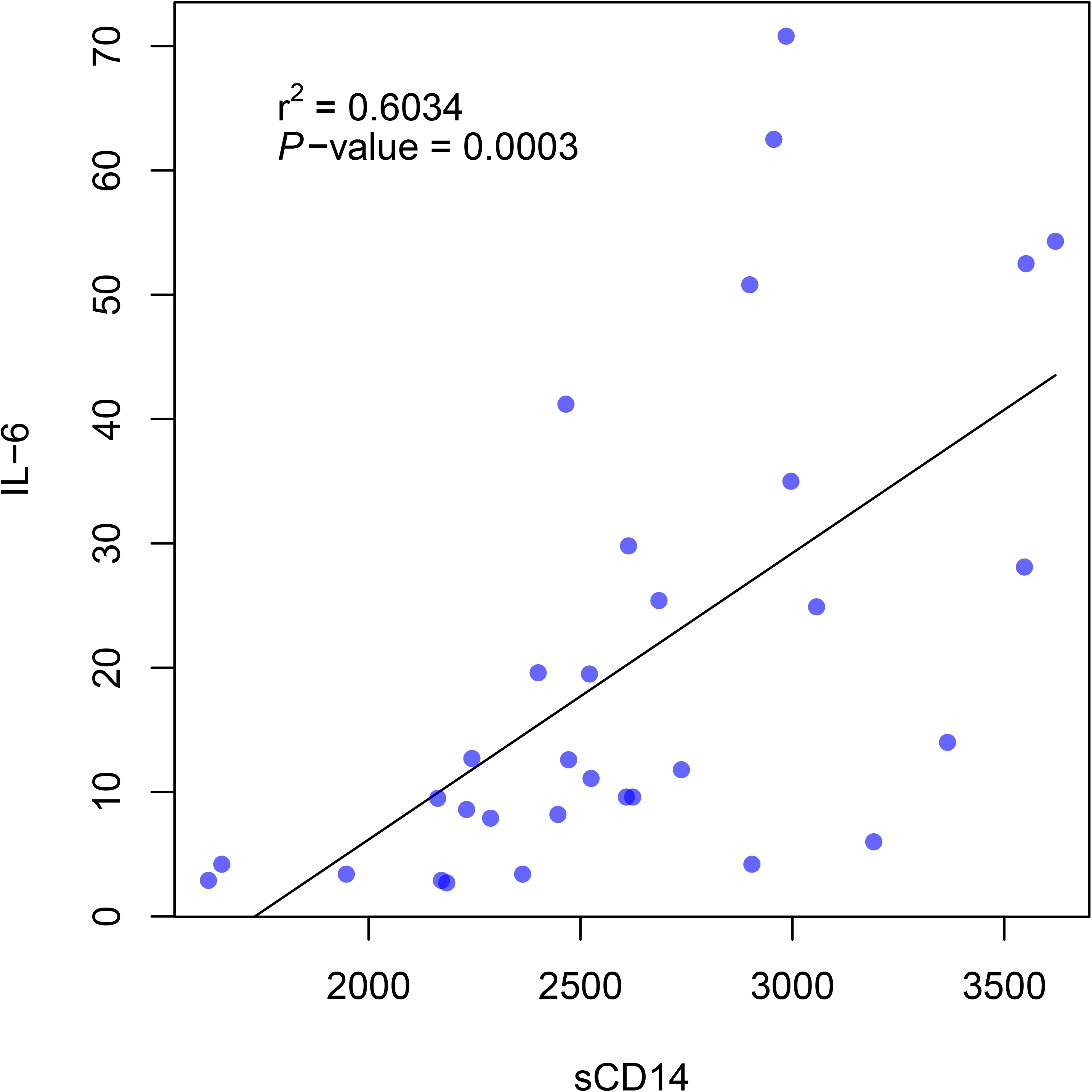
Association between serum levels of sCD14 and IL-6 levels in the non-ICU patient group. Pearson’s correlation coefficient (r^2^) and *P*-value are presented.

### Age-dependence of sCD14 and sCD163 levels

We analyzed possible age-dependence of sCD14 and sCD163 levels. Values did not show association between these biomarker levels and the age of patients.

## Discussion

Our results show, for the first time, increased levels of sCD14 and sCD163 in sera from SARS-Cov-2 infected patients admitted to hospital. We did not observe any differences between ICU or non-ICU patients, probably due to the interference on monocyte function produced by the use of tocilizumab and/or corticoid treatment in ICU patients as previously demonstrated [18; 19]. However, levels of sCD14 showed a strong correlation with clinical laboratory parameters, including acute phase reactants (ferritin, LDH, C-reactive protein, procalcitonin) and a strong correlation with IL-6 levels in the non-ICU patient group, where no tocilizumab and/or corticoids treatments were used. Furthermore, sCD163 levels showed a correlation with the time elapsed from hospital admission to sample extraction, increasing its value over time of hospital admission, suggesting a potential indicator of progression of disease.

Monocytes and macrophages constitute a key component of immune responses against viruses, acting as bridge between innate and adaptive immunity [20]. Activation of macrophages has been demonstrated to be pivotal in the pathogenesis of the immunosuppression associated to several viral infections (VIH, measles), where expansion of specific subsets of monocytes and macrophages in peripheral blood are observed, and considered to be drivers of immunopathogenesis [21]. Our results support the hypothesis of a preponderant role for monocytes in SARS-Cov-2 immunopathology, associated to an over exuberant immune response. Increased levels of monocyte-macrophage activation markers and the correlation with other inflammatory biomarkers (particularly IL-6), indicate a close relationship between monocyte activation and immunopathology in these patients. Inflammatory markers are closely related to severity in COVID-19 pathology [22] and selective blockade of IL-6 has been demonstrated to be a good therapeutic strategy in COVID-19 pathology [23]. Our results thus suggest that monocyte-macrophage activation can act as driver cells of the cytokine storm and immunopathology associated to severe clinical course of COVID-19 patients. Further, monitorization of monocyte activity trough these soluble activation markers and/or follow-up of circulating inflammatory monocytes in peripheral blood, could be useful to assess disease progression in the same way as in other viral infections [16].

In addition, our results identify monocyte-macrophage as a good target for the design of therapeutic intervention using drugs that inhibit monocyte-macrophage activation and differentiation. In this sense, anti-GM CSF inhibitor drugs, currently under clinical trials for rheumatic and other auto-inflammatory diseases, might provide satisfactory results in COVID-19 patients. Other drugs targeting monocyte and/or macrophage could also be useful in COVID-19, as in other inflammatory diseases [24]. The strategy of inhibiting monocyte differentiation has proved useful in avoiding cytokine storm syndrome after CAR-T cell immunotherapy [25], suggesting a possible therapeutic application to COVID-19 immunopathology [26] The present study has several limitations, including the relatively low numbers of patients tested and the interference of tocilizumab and corticoids in ICU patients’ results. However, these preliminary results are strongly suggestive of an important implication of monocyte-macrophage in COVID-19 immunopathology, as highlighted by the correlations found between these biomarker levels and inflammatory parameters. Further studies using broader series are needed to confirm our findings.

In summary, our data underscore the preponderant role of monocyte and macrophage immune response in COVID-19 immunopathology and provide pointers for future interventions in drug strategies and monitoring plans for these patients.

## Data Availability

All data referred to this manuscript are available upon request

## Funding

This study received support from the Instituto de Salud Carlos III: project GePEM (Instituto de Salud Carlos III(ISCIII)/PI16/01478/Cofinanciado FEDER), DIAVIR (Instituto de Salud Carlos III(ISCIII)/DTS19/00049/Cofinanciado FEDER; Proyecto de Desarrollo Tecnológico en Salud) and Resvi-Omics (Instituto de Salud Carlos III(ISCIII)/PI19/01039/Cofinanciado FEDER) and project BI-BACVIR (PRIS-3; Agencia de Conocimiento en Salud (ACIS)—Servicio Gallego de Salud (SERGAS)—Xunta de Galicia; Spain) given to A.S.; and projects ReSVinext (Instituto de Salud Carlos III(ISCIII)/PI16/01569/Cofinanciado FEDER), and Enterogen (Instituto de Salud Carlos III(ISCIII)/ PI19/01090/Cofinanciado FEDER) given to F.M.-T.

We want to acknowledge the effort of all first-line healthcare workers of the patients included in this study.

